# Approaches to handle missing follow-up time: A comparative analysis of contralateral breast cancer incidence

**DOI:** 10.1101/2025.05.28.25327971

**Authors:** Sarah R Haile, Miriam Wanner, Dimitri Korol, Sabine Rohrmann

## Abstract

**Background:** We aimed to compare various common approaches for handling missing vital status or follow-up time. As a case study for application of these methods, we estimated incidence of metachronous contralateral breast cancer (CBC).

**Methods:** For 1980-2016, incidence of metachronous CBC with follow-up through 2024 was estimated using Poisson regression with overdispersion, by age at incidence, year of incidence, histology and follow-up period. Missing follow-up time was ignored in the naive approach, simulated once using the average hazard derived from published Swiss cancer registry data, or multiply imputed using 3 different imputation models.

**Results:** 24,612 women aged 20-84 had unilateral breast cancer between 1980 and 2016 in the Swiss cantons of Zurich and Zug. Of those, 5% (n=1264) were lost to follow-up. Over 291463 person-years, 1145 contralateral breast malignancies were diagnosed, corresponding to 393 per 100000 person-years (95% CI 353 to 438). Incidence rates have been decreasing over time to 238 (171 to 333) for the incidence period 2010-2016. The same overall pattern was observed regardless of how we handled missing follow-up times. However, using a single imputation generally produced lower incidence rates compared to the naive approach, with multiple imputation giving higher estimates. The most complex multiple imputation model gave incidence estimates that were very similar to those from the naive approach.

**Conclusion:** Different methods to handle missing follow-up times yielded similar results: that CBC incidence has declined in recent decades. Multiple imputation is likely an appropriate method to handle missing follow-up data, enabling researchers to include all eligible individuals in the analysis.

## Introduction

Data collected by cancer registries is necessary to understand which cancers are diagnosed at what frequency, and what happens with people with cancer after the initial diagnosis. Such data aids in answering questions such as how effective cancer screening programs are, and which individuals are likely to be diagnosed with a second primary tumor. To accomplish these goals, cancer registries must ensure the data they collect is accurate and up-to-date. Well-established guidelines on data quality in cancer registries recommend assessing four key indicators: comparability, validity, timeliness, and completeness. Comparability assesses whether data collection procedures follow international guidelines, such as those outlined by the European Network of Cancer Registries (ENCR) (European Network of Cancer Registries 2025). Validity examines whether diagnoses were morphologically verified. Timeliness looks at in what time frame cancer diagnoses can expect to be available to the registry and later in the registry’s database. Finally, Completeness assesses what proportion of all incident cancers in the corresponding population are reported to the registry (Bray and Parkin 2009; Parkin and Bray 2009). In Switzerland, the Cancer Registry Zurich and Zug has previously reported on these indicators (Wanner et al. 2018).

One aspect of comparability is certainly having accurate and up-to-date follow-up times and if the patient has died, a date of death (Bray and Parkin 2009, Table 1). This information is especially important for estimating not only absolute and relative overall survival (Brenner and Hakulinen 2009), but also incidence of e.g. second primary cancers. Obtaining correct vital status and date of death varies greatly according to the legal and administrative situation, as it often requires linkage to vital statistics offices. If a unique identifier is not used, such linkage may depend on probability matching. Sometimes death of registered patients has been missed due to incorrect linkage or emigration out of the registry’s geographic area, or missing ability of the registry to conduct a (full) linkage, leading to “immortals”, that is, individuals that appear to be alive forever in the registry as death information can no longer be obtained (Andersson et al. 2022).

**Table 1:**
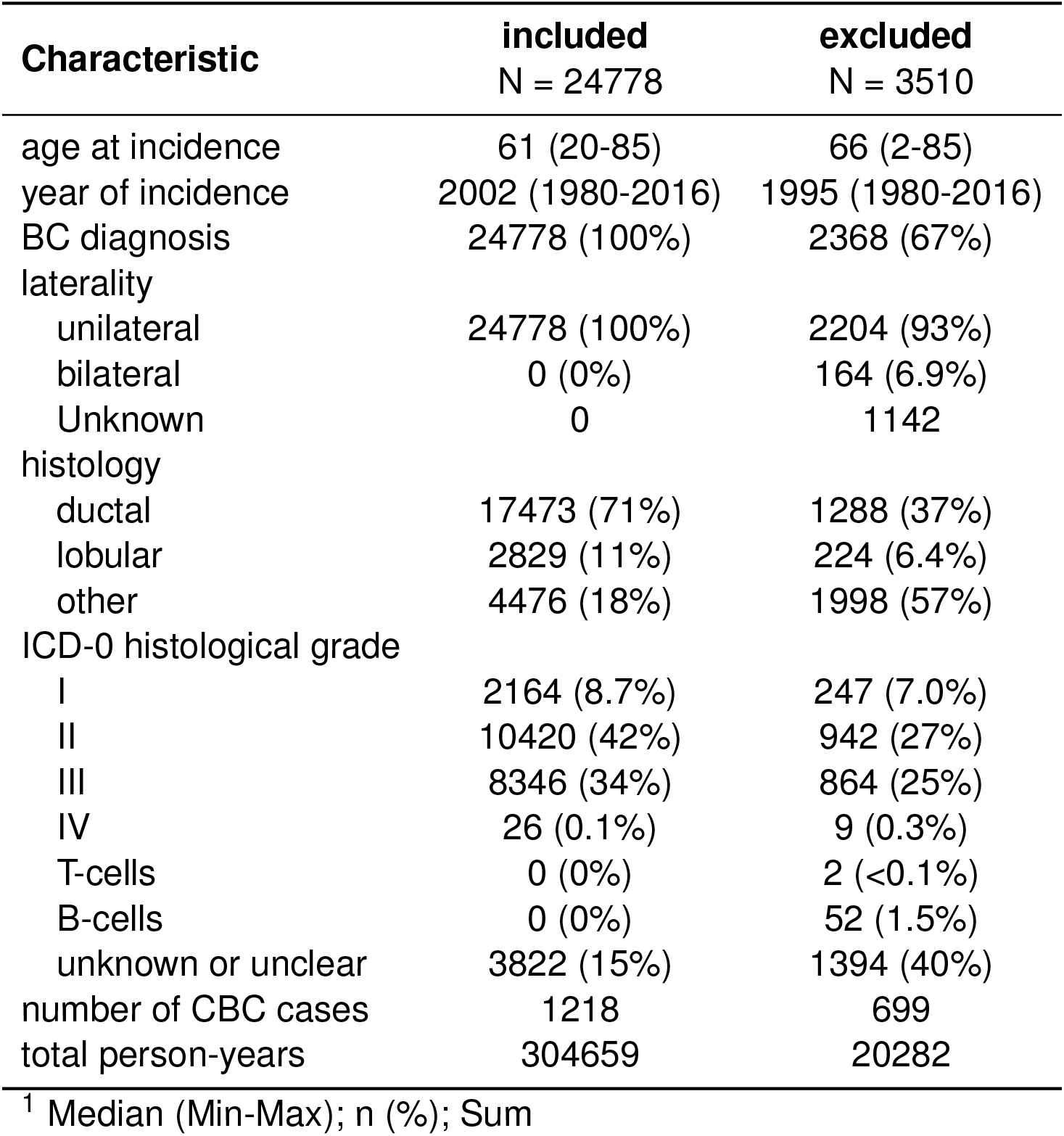
Characteristics of included and excluded participants in the cancer registries from the Swiss cantons of Zurich and Zug.

While the situation in Switzerland with respect to such immortal participants has improved for newly registered patients due to the 2020 Cancer Registration Act (Krebsregistrierungsgesetz (KRG) 2023), some older cases remain immortal in the registry due to lack of linkage possibilities and lack of digitized data in earlier years. It is not always clear how to analyze such data. A variety of *ad hoc* methods have been used in analyses of cancer registry data to handle individuals with missing follow-up time, missing vital status or potentially immortal status. While some publications that have likely presented such data do not mention handling of missing data, others use a so-called simulation approach, creating a single imputation based on hazard rates from cancer registry data. Previously, we performed a simulation study assessing how 3 approaches (ignoring missing follow-up times and vital status, single imputation or multiple imputation (Sterne et al. 2011)) worked in typical analysis tasks of a cancer registry where the true value of the outcome was known, and came to the conclusion that multiple imputation was likely the least biased approach (Richter et al. 2024).

In this analysis, we aimed to determine the best statistical approach to handle individuals with missing follow-up or vital status in a common cancer registry analysis where the true value is not known. Therefore we updated an 2016 analysis of metachronous contralateral breast cancer incidence (Prater et al. 2016), and compared the results after adjusting for missing vital status or follow-up time in different ways.

## Methods

The epidemiological Cancer Registry of the Cantons of Zurich, Zug, Schaffhausen and Schwyz has been been collecting data on cancer cases in the Swiss canton of Zurich since 1980, in the canton of Zug since 2011 and in the cantons of Schaffhausen and Schwyz since 2020. Included in this analysis are the two cantons, Zurich and Zug, that provide at least 5 years of follow-up. Together, Zurich and Zug have a population of around 1.7 million inhabitants and make up about 20% of the total population of Switzerland (Bundesamt für Statistik 2024).

This analysis included all female patients with a unilateral breast cancer from the cantons of Zurich or Zug in the time period from 1980 to 2016, with follow-up until at least 2021. Of the 28,288 patients identified in the registry, 7 were excluded as either younger than 20 or older than 85 years. A further 1135 were excluded because of other malignancies prior to the first breast cancer incidence. To distinguish between synchronous and metachronous cancers, we excluded 2229 participants with follow-up for 90 days or shorter. Finally, 305 subjects were excluded because of missing laterality or a first bilateral breast cancer.

The person-time at risk for the calculation of incidence of contralateral breast cancer (CBC) was counted from 90 days after diagnosis of the initial primary breast cancer until occurrence of CBC or censored at death or end of follow-up period. Quasi-poisson regression to account for overdispersion was performed to estimate incidence rates and incidence rate ratios of metachronous CBC stratified by age at diagnosis (5-year categories) and incidence year (possibly grouped into 5-year categories) of unilateral breast cancer (UBC, in 5-year periods), histology of UBC (ductal, lobular, other) and follow-up period (3-year intervals). Results were presented as number of cases per 100,000 person-years, and corresponding 95% confidence intervals (95% CI) were obtained.

Several different pragmatic approaches were considered to fill in the missing follow-up times, that is, cases where the cancer registry have denoted the status as “lost to follow-up” or where the most recent update of vital status was several years ago. In the naive approach (NI), the person-years were left as they were in the original data. In the single imputation approach (SI), person-years for individuals who were lost to follow-up but did not have a CBC were simulated once with Swiss breast cancer survival data (publicly available (National Agency for Cancer Registration 2024)). A constant hazard rate of 0.0591 per year for survival after UBC was assumed for the whole period from 1980 to 2021 to compute random survival time, similar to the 0.0591 used in a previous analysis of CBC in this population (Prater et al. 2016). This method was included as it seems commonly used for this kind of missing data in cancer registries, even though it may introduce bias since not every patient could be assumed to have the same hazard rate in such a heterogenous population. Additionally 3 multiple imputation (MI) approaches were considered. MI model A adjusted for age group, incidence period, morphology and the Nelson-Aalen estimator of the baseline hazard (White and Royston 2009). Considering age and incidence period as continuous variables, MI model B included age (centered at 60), age-squared (centered at 60), incidence year (centered at 2001), incidence year squared (centered at 2001), morphology and baseline hazard. The simplest model, MI model C, included age group, incidence period and morphology only. These MI models were chosen to be typical options that might be used in practice, for example to handle non-linear effects of age or incidence year, or to handle survival outcomes, and are not intended to be an exhaustive set of options. The 3 MI approaches created 10 imputations each and incidence estimates were pooled using Rubin’s rules (Rubin 1987). Predictive mean matching was used to impute all variables.

Statistical analyses were performed in R (version 4.5.1 (R Core Team 2024)). Multiple imputation was performed using the mice package (van Buuren and Groothuis-Oudshoorn 2011).

## Results

A total of 24778 women with UBC aged 20-84 years met the inclusion criteria for the present analysis. Patient characteristics are summarized in Table 1. Of the included women, 5.1% (n = 1256) were defined by the cancer registry as being lost to follow-up or had no follow-up after 2021. Missingness rates by key variables are found in Table S1.

Over 304659 person-years, 1218 contralateral breast malignancies were diagnosed, corresponding to an overall incidence rate of 415 per 100,000 person years (95% CI 374 to 460) (Table 2, MI (B) approach). (In this paragraph, we focus on the CBC incidence rates estimating using the MI (B) approach, while methodological comparisons can be found in the following paragraph.) Incidence rates decrease somewhat with age, ranging from 512 (350 to 750) for those diagnosed in their 30s down to 325 (190 to 556) for those diagnoses after the age of 80. Recent decades have also seen a steady decrease in CBC incidence, estimated to be 244 per 100,000 (176 to 340) for those diagnosed between 2010 and 2016. Examining the incidence rates by histology, those with initial ductal histology had the lowest incidence of CBC (394, 347 to 447). Incidence rates were fairly constant over the follow-up period (Figure S1 and Table S2).

**Table 2:**
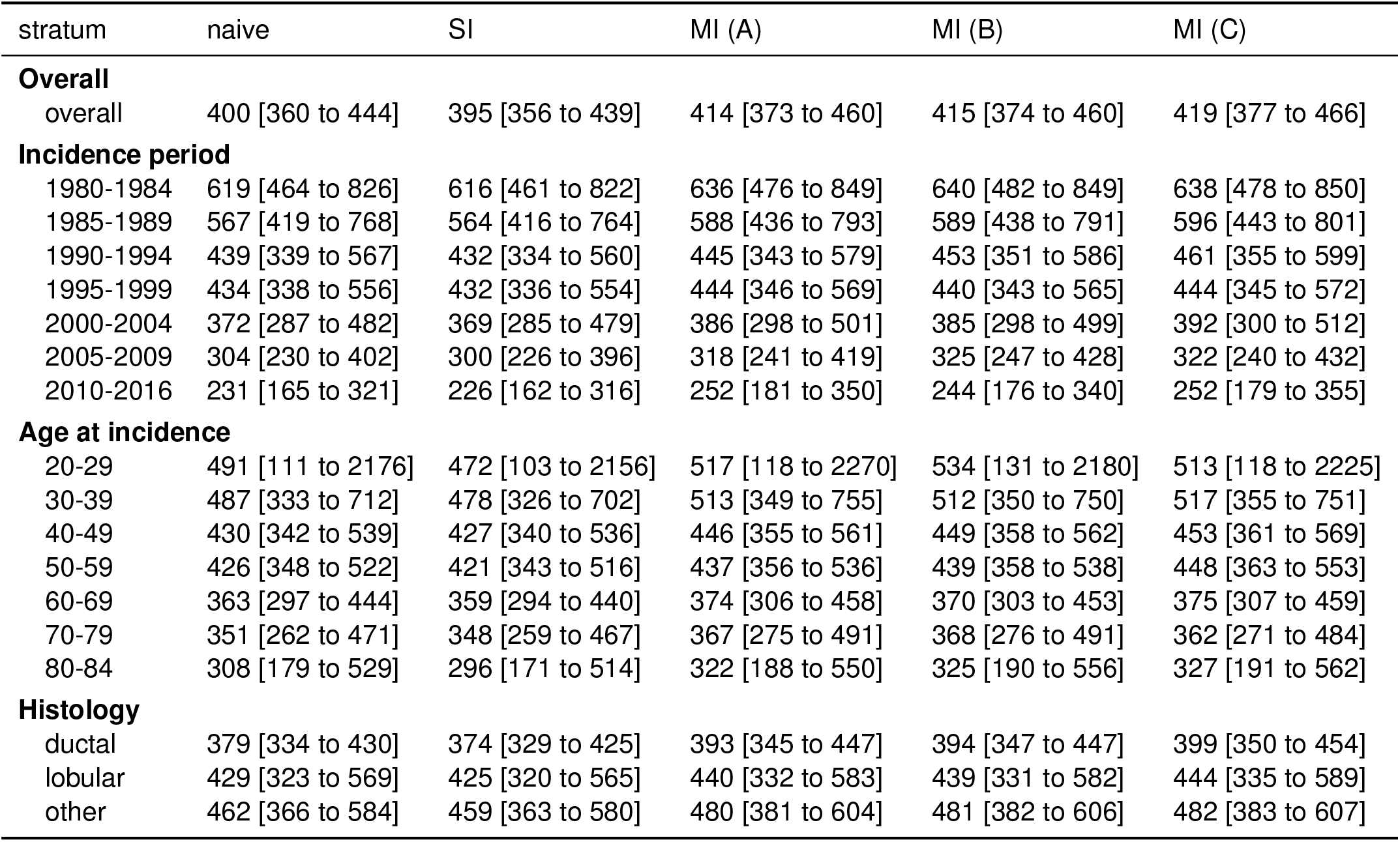
estimated CBC incidence per 100,000 person years, by incidence period, age category, and histology as well as by approach to handle missing follow-up time.

Comparing the results by method used to handle missing follow-up times, we can examine both the magnitude of the incidence estimates, as well as the width of the corresponding 95% CIs. The naive approach tended to be 10 to 20 cases per 100,000 person years lower than the other approaches (Table 2 and Figure 2). SI produced the lowest estimates of CBC incidence in all subgroups. The MI models A and C tended to have lowerer incidence estimates than model B, which appeared closest to the average subgroup incidence. Regarding CI width, SI produced narrower CIs in all subgroups, while MI models A and C generally produced the widest CIs (Table 2 and Figure 3).

**Figure 1:**
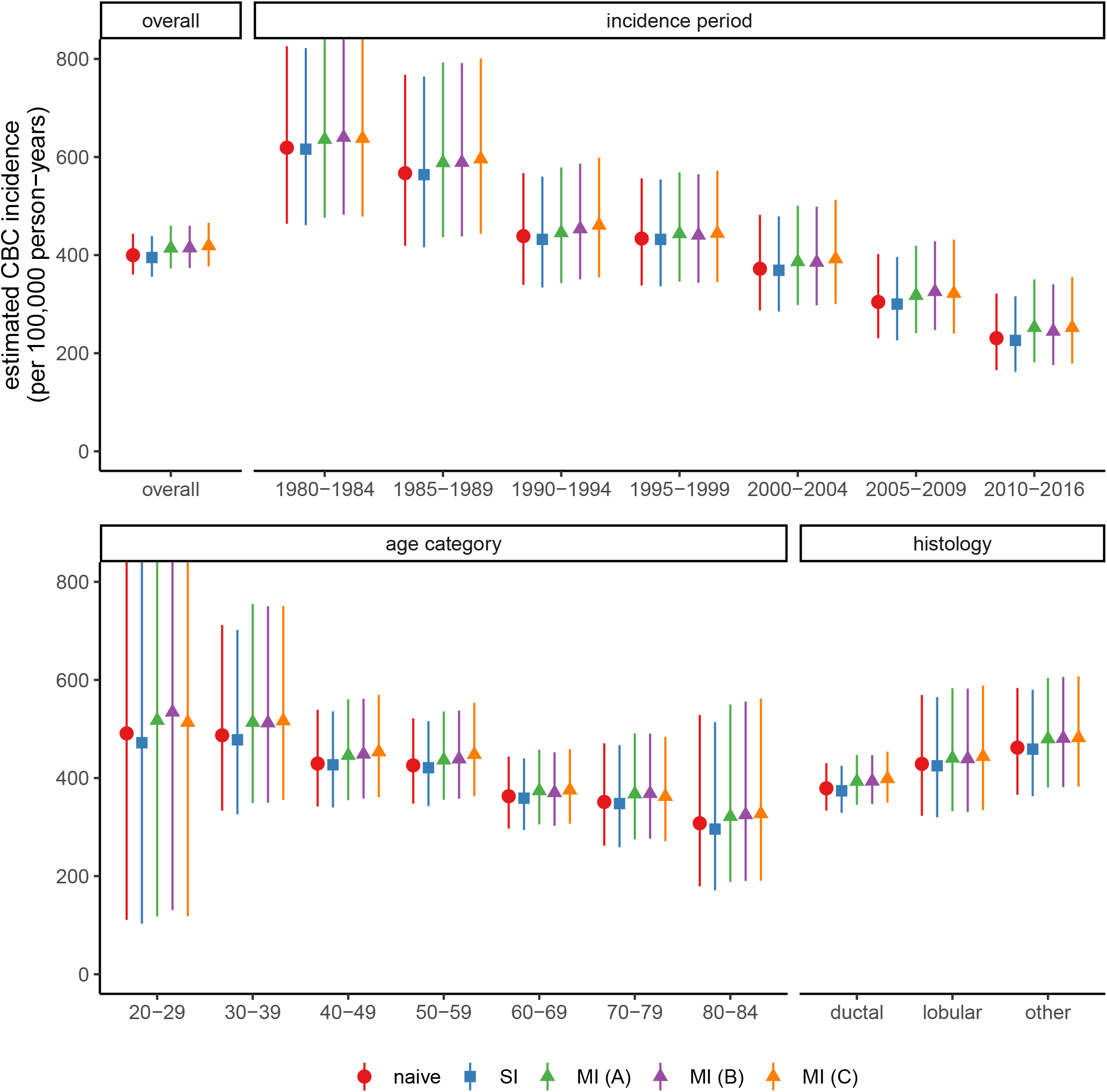
Estimated CBC incidence per 100,000 person-years, overall, and by incidence period, age category and histology. Primarily, MI (B, in purple) estimates are discussed in the text.

**Figure 2:**
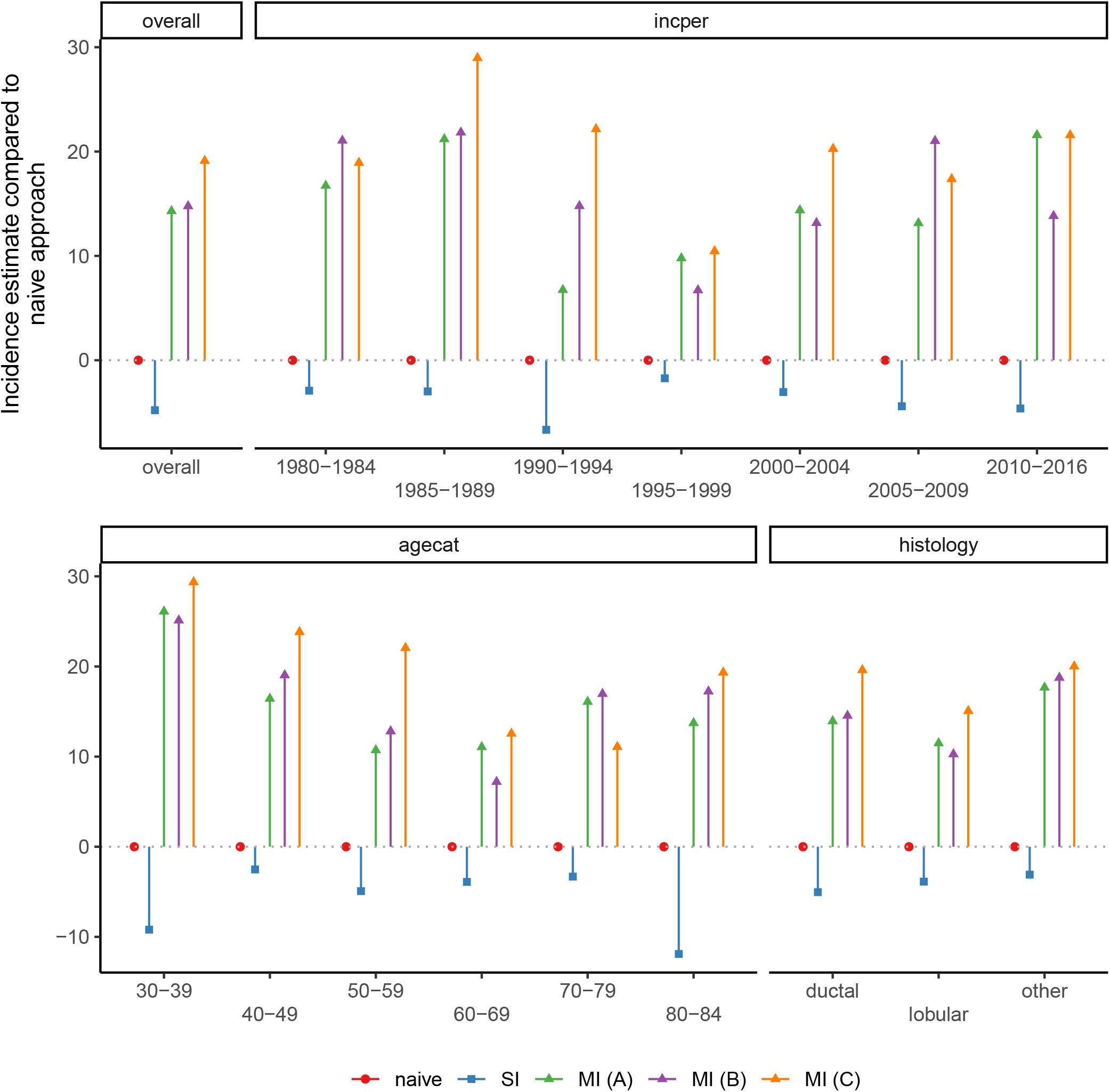
Comparison of CBC incidence estimates by method for handling missing follow-up times. Naive, SI and all MI estimates are compared to the naive approach.

**Figure 3:**
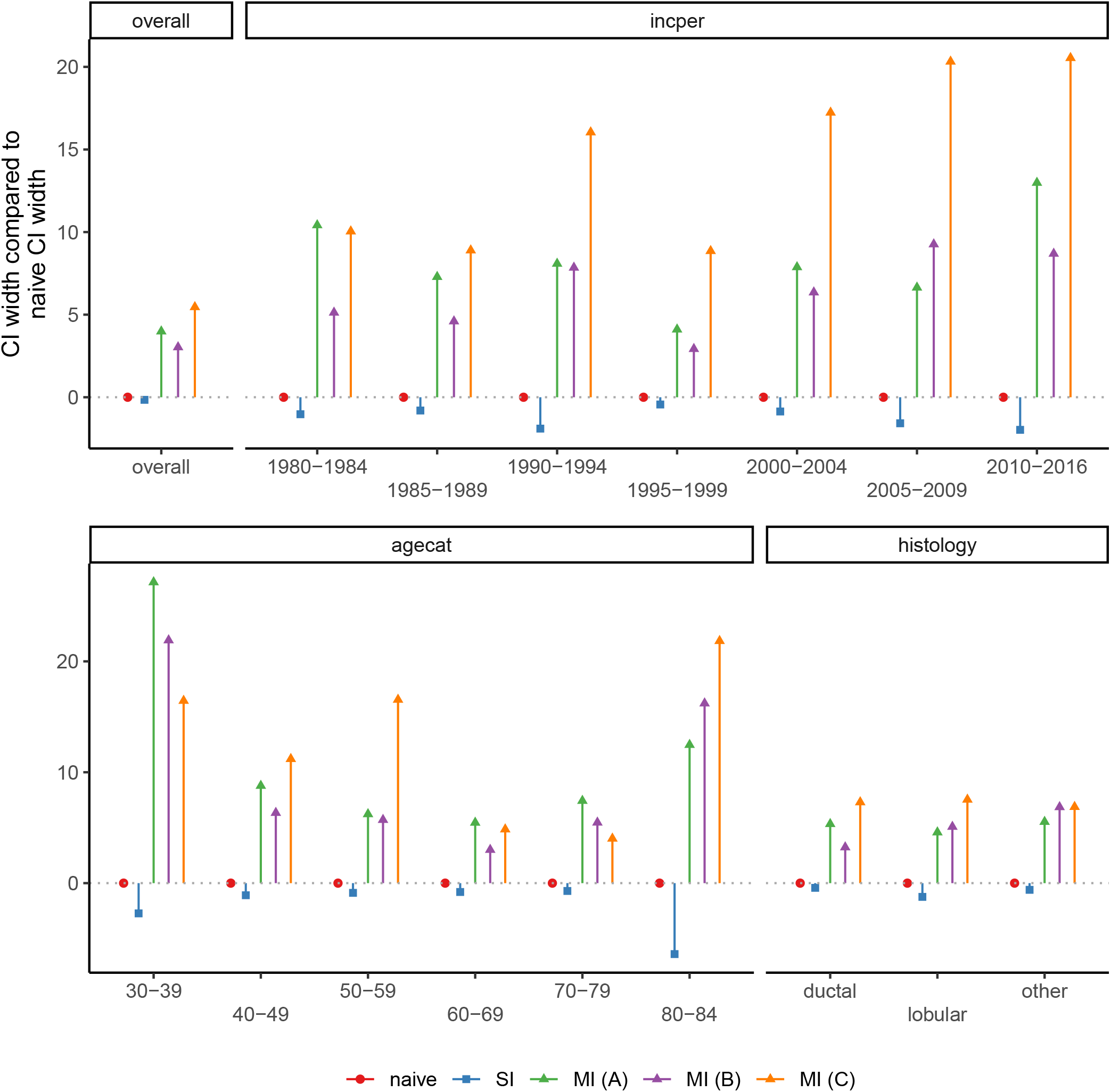
Comparison of width of confidence interval for CBC incidence estimates by method for handling missing follow-up times. SI and all MI estimates are compared to the naive approach within each subgroup.

## Discussion

Regardless of which method was used to deal with missing follow-up or vital status, incidence of metachronous CBC has decreased in recent years, following trends described by e.g. Prater et al. (2016). This decrease in incidence appears to be mostly driven by lower CBC incidence for those women with first BC incidence in recent years, a pattern likely due to the increased use of hormone therapies such as tamoxifen (Early Breast Cancer Trialists’ Collaborative Group (EBCTCG) 2011; Nordenskjöld et al. 2023). Younger patients continue to have a somewhat higher risk of CBC than older patients, and those with ductal or lobular histology have lower risk than those with other histologies.

With respect to the choice of approach for handling missing follow-up or vital status, the various approaches yielded by and large similar results. CBC incidence estimates from SI models were generally smaller than those of other methods and had narrower confidence intervals, an expected phenomenon of SI as it produces standard errors which are too small (Sterne et al. 2011). MI (regardless of choice of imputation model) produced estimates of CBC incidence that were generally within ± 10 units of the naive approach, but with slightly wider confidence intervals to account for the missing follow-up times. While patients with missing follow-up times could be excluded from the analysis when using the naive approach, MI enables them to be included, and permits analysis with other missing covariates as well. We recommend using MI therefore in all cases where missing vital status, missing follow-up time, or other missing covariate information is present to enable data from all patients to be included in the statistical analysis. The naive approach should only be used if there is no danger of “immortal” patients and if patients will not be unnecessarily excluded due to missing covariates.

Missing covariate information can lead to exclusion of patients from the statistical analysis, leading to biased estimates of the outcome of interest without further adjustment such as MI (Pitcha et al. 2023). Using MI to account for missing covariates is not new in analysis of cancer registry, however it is often used to fill in missing baseline characteristics such as tumor stage or hormone receptor status (Nur et al. 2010; Eisemann et al. 2011; Howlader et al. 2012; Luo et al. 2017; Parry et al. 2019). It is well-known that missing vital status and missing event times can be imputed (White and Royston 2009; Carpenter et al. 2023). However, we are unaware of assessments of the use of MI in cases where vital status or follow-up date are missing, with the exception of our previous simulation study (Richter et al. 2024). In that simulation study we concluded that MI produced least biased results in a number of typical tasks of cancer registries, such as overall survival, relative survival and standardized incidence ratio. Incorporating MI into otherwise straightforward analysis tasks gives us reduced bias in the estimated outcomes in exchange for somewhat more complex statistical programming. In Richter et al. (2024), we outlined R and Stata code to perform survival analysis after imputation of missing follow-up times. The *online resource* here outlines R and Stata code to estimate incidence rates in a similar setting.

The main strengths of this analysis are the large dataset from a cancer registry with up to 35 years of follow-up (median 10 years), and the prespecified analysis plan with many potential approaches to handle missing follow-up data. We were able to replicate CBC incidence rates from a previous CBC analysis (Prater et al. 2016), without having the previous version of the data. A key limitation is that the true CBC incidence rate is not known, so we could not compare the results of the missing data approaches to a true value. We are aware that we do not have all variables associated with missingness in vital status or follow-up time, but we believe we have the main ones. Additionally, we did not consider other potential predictors of CBC, such as hormone therapy, type of surgery, use of radiation, lymph node involvement, time from diagnosis to treatment, or genetic predisposition (Akdeniz et al. 2019; Syleouni et al. 2023). However, predictors of missingness in baseline covariates such as those just listed are not generally good predictors of missingness in vital status in cancer registries.

## Conclusions

Incidence rates of CBC have been in steady decline in Switzerland since the 1980s, a trend seen in other studies and likely due to increased use of hormone therapies such as tamoxifen. Multiple imputation can be used to fill in missing follow-up times or vital status, enabling inclusion of all eligible individuals in the analysis. Its use would improve data quality in analyses of cancer registry data.

## Statements & Declarations

### Author contributions

Study concept and design was developed by SH and SR. Data collection was performed by MW and DK. Statistical analysis was performed by SH. The first draft of the manuscript was written by SH, and all authors provided feedback on the manuscript prior to submission. All authors read and approved the final manuscript.

### Data availability

Data analyzed in the current study are not publicly available. A request for anonymized data can be submitted to the National Agency for Cancer Registration. The request procedure is described at https://nacr.ch/en/information-for-researchers/request-for-anonymised-data.

### Ethics approval

This is a data quality study of previously collected cancer registry data. No ethical approval is required.

### Consent to participate

Patients with new cancer diagnoses in Switzerland have the right to information about cancer registration, have the right to view their data, and have the right to object to registration (https://nacr.ch/en/information-for-patients). Such an objection can be submitted or withdrawn at any time, and is valid throughout the patient’s life and across Switzerland. Personal details, such as name, address, date of birth, and social insurance number are not forwarded to the NACR.

### Consent to publish

The manuscript contains no individual patient data.

