## Supplemental Table for "Approaches to handle missing follow-up time: A comparative analysis of contralateral breast cancer incidence"

### Contents


**Predictors of missingness** **2**

**Comparisons of results by follow-up period** **3**

**Code to multiply impute follow-up times and vital status** **4**

R code . . . . . . . . . . . . . . . . . . . . . . . . . . . . . . . . . . . . . . . . . . . 4

Stata code . . . . . . . . . . . . . . . . . . . . . . . . . . . . . . . . . . . . . . . . . 6

**Computational Details** **9**

#### Predictors of missingness

**Table S1:** Proportion of missing follow-up by age at first incidence, incidence period, histology and follow-up period

| **Characteristic** | **lost to follow-up** N = 1,256 | **up to date** N = 23,522 |
|---|---|---|
| age at first incidence | | |
| 20-29 | 9 (7.0%) | 119 (93.0%) |
| 30-39 | 130 (9.6%) | 1,220 (90.4%) |
| 40-49 | 358 (7.7%) | 4,295 (92.3%) |
| 50-59 | 366 (6.3%) | 5,489 (93.7%) |
| 60-69 | 211 (3.4%) | 6,013 (96.6%) |
| 70-79 | 136 (2.8%) | 4,751 (97.2%) |
| 80-84 | 46 (2.7%) | 1,635 (97.3%) |
| incidence period | | |
| 1980-1984 | 87 (3.8%) | 2,186 (96.2%) |
| 1985-1989 | 133 (5.3%) | 2,391 (94.7%) |
| 1990-1994 | 153 (5.7%) | 2,547 (94.3%) |
| 1995-1999 | 127 (4.1%) | 3,005 (95.9%) |
| 2000-2004 | 189 (5.0%) | 3,568 (95.0%) |
| 2005-2009 | 238 (6.0%) | 3,731 (94.0%) |
| 2010-2016 | 329 (5.1%) | 6,094 (94.9%) |
| histology | | |
| ductal | 922 (5.3%) | 16,551 (94.7%) |
| lobular | 117 (4.1%) | 2,712 (95.9%) |
| other | 217 (4.8%) | 4,259 (95.2%) |
| follow-up period | | |
| 0-4 | 455 (8.3%) | 5,044 (91.7%) |
| 5-9 | 422 (6.9%) | 5,662 (93.1%) |
| 10-14 | 200 (3.8%) | 5,011 (96.2%) |
| 15-19 | 90 (2.6%) | 3,334 (97.4%) |
| 20-29 | 73 (2.1%) | 3,415 (97.9%) |
| 30-44 | 16 (1.5%) | 1,056 (98.5%) |

[1] n (%)

#### Comparisons of results by follow-up period

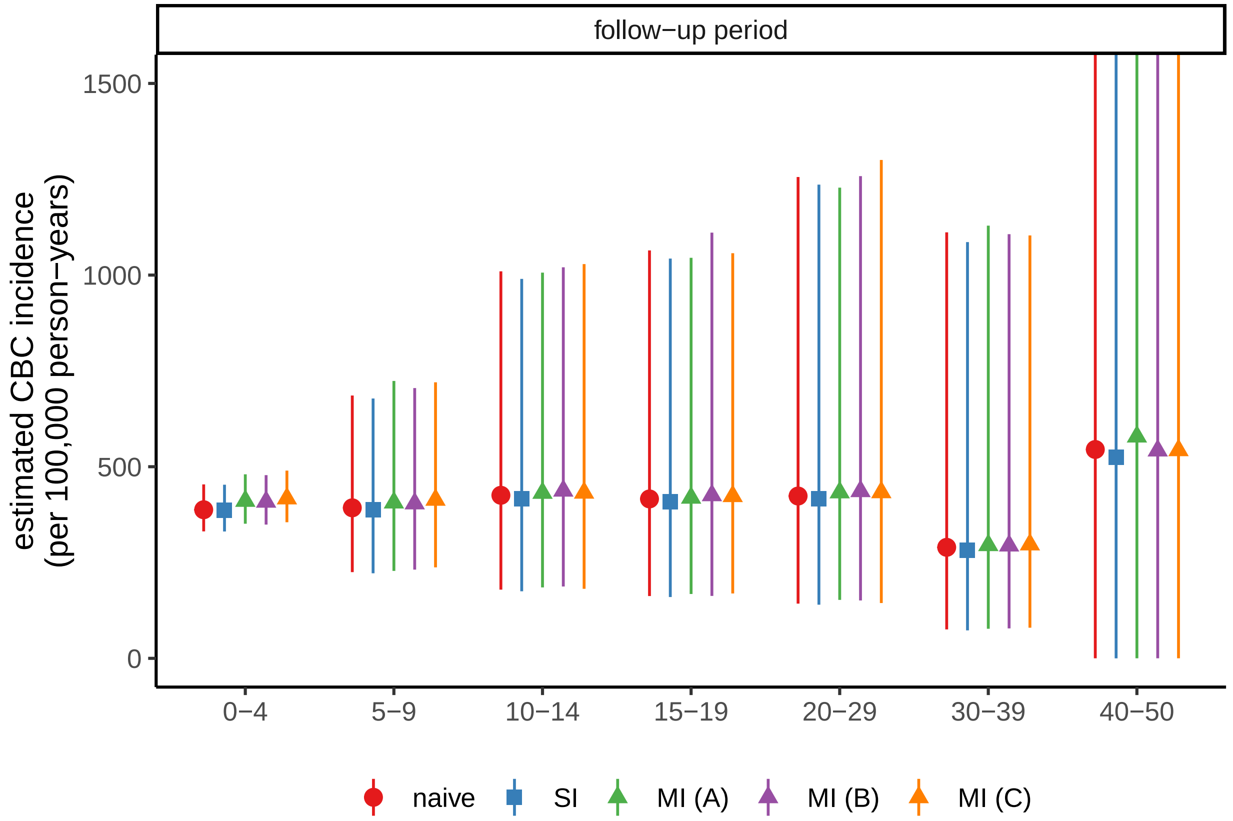


**Figure S1:** Comparison of estimated CBC incidence, by follow-up period

**Table S2:** estimated CBC incidence per 100,000 person years, by follow-up as well as by approach to handle missing follow-up time

| stratum | naive | SI | MI (A) | MI (B) | MI (C) |
|---|---|---|---|---|---|
| **Follow-up period** | | | | | |
| 0-4 | 388 [331 to 454] | 387 [331 to 453] | 411 [351 to 480] | 408 [349 to 478] | 417 [355 to 490] |
| 5-9 | 393 [225 to 686] | 388 [222 to 678] | 406 [228 to 724] | 404 [232 to 705] | 414 [237 to 720] |
| 10-14 | 425 [179 to 1010] | 417 [175 to 990] | 432 [185 to 1006] | 437 [187 to 1020] | 432 [181 to 1029] |
| 15-19 | 416 [163 to 1064] | 408 [160 to 1043] | 419 [168 to 1045] | 425 [163 to 1111] | 423 [169 to 1057] |
| 20-29 | 424 [143 to 1256] | 417 [140 to 1236] | 433 [153 to 1228] | 436 [151 to 1258] | 433 [144 to 1300] |
| 30-39 | 290 [75 to 1112] | 282 [73 to 1086] | 295 [77 to 1129] | 294 [78 to 1107] | 297 [80 to 1103] |
| 40-50 | 545 [0 to 2326767] | 525 [0 to 2269161] | 578 [0 to 1812224] | 542 [0 to 2299532] | 543 [0 to 2313425] |

#### Code to multiply impute follow-up times and vital status

##### R code

This R code fits poisson models with overdispersion using the `quasipoisson` family in the `glm()` regression function. Alternatively, poisson models can be fit using the `poisson` family, or negative binomal regression using `glm.nb()` available from the `MASS` package. We have computed incidence rates for each of the imputed datasets with `glht` from the `multcomp` packages and then pooled the coefficients. The reverse should in theory be possible, but `glht` does not accept pooled model results from `mice`.

```
library(mice)
library(multcomp)

# if subjects are defined as lost to follow-up, set
# event time and vital status to missing
small_dat <- testdata %>%
  mutate(t = ifelse(is_lfup, NA, t),
         e = ifelse(is_lfup & e != 1, NA, e)) %>%
  mutate(agec = agei - 60, # centered age
         age2 = agec ^ 2, # age^2
         yyc = yyi - 2001, # centered year of incidence
         yy2 = yyc ^ 2, # year of incidence ^ 2
         ) %>%
  dplyr::select(-agei, -yyi, -is_lfup)

head(small_dat)
```

| histology | t | e | agec | age2 | yyc | yy2 |
|---|---|---|---|---|---|---|
| ductal | 26.503593 | 0 | -14.674881 | 215.35213 | -9 | 81 |
| ductal | 16.336927 | 0 | 5.607117 | 31.43976 | -9 | 81 |
| ductal | 31.586927 | 0 | -5.056812 | 25.57135 | -9 | 81 |
| ductal | 10.086927 | 0 | -9.084190 | 82.52251 | -9 | 81 |
| ductal | 4.253593 | 1 | -20.829567 | 433.87086 | -9 | 81 |
| ductal | 31.503593 | 0 | -8.284737 | 68.63686 | -9 | 81 |

```r
imp <- mice(small_dat, m = 20, printFlag = FALSE,
            seed = 123456)
mod <- with(imp, glm(e ~ 1 + offset(log(t)),
                 family = quasipoisson))
summary(pool(mod), conf.int = TRUE) %>%
    mutate(incid = exp(estimate),
         lb = exp(`2.5 %`),
         ub = exp(`97.5 %`)) %>%
  dplyr::select(term, estimate, `2.5 %`, `97.5 %`,
                incid, lb, ub) %>%
  kable(digits = c(1, 2, 2, 2, 6, 6, 6))
```

| term | estimate | 2.5 % | 97.5 % | incid | lb | ub |
|---|---|---|---|---|---|---|
| (Intercept) | -5.48 | -5.59 | -5.38 | 0.004154 | 0.00375 | 0.0046 |

```r
mod <- with(imp, glm(e ~ histology + offset(log(t)),
                 family = quasipoisson))

mcs <- map(mod$analyses, ~ glht(., linfct = c("(Intercept) = 0",
                                             "(Intercept) + histologylobular = 0",
                                             "(Intercept) + histologyother = 0")))
pool(mcs)$pooled %>%
  mutate(`2.5 %` = estimate - 1.96 * sqrt(t),
         `97.5 %` = estimate + 1.96 * sqrt(t)) %>%
  mutate(incid = exp(estimate),
         lb = exp(`2.5 %`),
         ub = exp(`97.5 %`)) %>%
  dplyr::select(term = contrast, estimate, `2.5 %`, `97.5 %`,
                incid, lb, ub) %>%
  mutate(term = c("ductal", "lobular", "other")) %>%
  kable(digits = c(1, 2, 2, 2, 6, 6, 6))
```

| term | estimate | 2.5 % | 97.5 % | incid | lb | ub |
|---|---|---|---|---|---|---|
| ductal | -5.53 | -5.66 | -5.41 | 0.003951 | 0.003488 | 0.004477 |

| term | estimate | 2.5 % | 97.5 % | incid | lb | ub |
|---|---|---|---|---|---|---|
| lobular | -5.43 | -5.73 | -5.13 | 0.004369 | 0.003240 | 0.005892 |
| other | -5.34 | -5.56 | -5.11 | 0.004807 | 0.003846 | 0.006008 |

#### Stata code

This stata code fits poisson models with overdispersion using the `glm` command with `family(poisson)` and `scale(x2)` options. Alternatively, the `poisson` command could be used directly if no adjustment for overdispersion is required (perhaps with the `vce(robust)` option for robust standard errors) or negative binomial regression fit with the `nbreg` command would also account for overdispersion. Stata pools the coefficients automatically in `mi estimate`, and its `post` option allows us to compute the incidence rates afterward with `lincom`.

```
##
## . version
## version 19.5
##
## . cd ~/
## /home/sahail
##
## . use testdata
##
## . replace t = . if is_lfup == 1
## (1,256 real changes made, 1,256 to missing)
##
## . replace e = . if is_lfup == 1 & e != 1
## (1,214 real changes made, 1,214 to missing)
##
## . gen agec = agei - 60
##
## . gen age2 = agec ^ 2
##
## . gen yyc = yyi - 2001
##
## . gen yy2 = yyc ^ 2
##
## . encode histology, gen(hist)
##
```

```
## .
## . set seed 123456
##
## . mi set mlong
##
## . mi register imputed t e
## (1256 m=0 obs now marked as incomplete)
##
## . qui: mi impute chained (pmm, knn(5)) t e = agec age2 yyc yy2 hist, add(20)
##
## .
## . mi estimate, post: glm e, exposure(t) family(poisson) scale(x2)
##
## Multiple-imputation estimates                    Imputations       =         20
## Generalized linear models                        Number of obs     =     24,778
##                                                  Average RVI       =     0.0161
##                                                  Largest FMI       =     0.0159
##                                                  DF:     min       =  75,842.40
##                                                          avg       =  75,842.40
## DF adjustment:   Large sample                           max       =  75,842.40
##                                                  F(   0,      .)   =          .
## Within VCE type:          OIM                    Prob > F          =          .
##
## ------------------------------------------------------------------------------
##            e | Coefficient  Std. err.      t    P>|t|     [95% conf. interval]
## -------------+----------------------------------------------------------------
##        _cons |  -5.488096   .0517592  -106.03   0.000    -5.589544   -5.386648
##        ln(t) |          1  (exposure)
## ------------------------------------------------------------------------------
##
## . lincom _cons, eform
##
##  ( 1)  [e]_cons = 0
##
## ------------------------------------------------------------------------------
##            e |     exp(b)   Std. err.      z    P>|z|     [95% conf. interval]
## -------------+----------------------------------------------------------------
##          (1) |   .0041357   .0002141  -106.03   0.000     .0037367    .0045773
## ------------------------------------------------------------------------------
##
## . di "overall incidence: ", r(estimate) * 100000, "[", r(lb) * 100000, ", ", r(
```

```
## > ub) * 100000. "]"
## overall incidence:  413.57109 [ 373.67377 ,  457.72826]
##
## .
## . mi estimate, post: glm e i.hist, exposure(t) family(poisson) scale(x2)
##
## Multiple-imputation estimates                  Imputations       =         20
## Generalized linear models                      Number of obs     =     24,778
##                                                Average RVI       =     0.0136
##                                                Largest FMI       =     0.0152
## DF adjustment:   Large sample                  DF:     min       =  82,366.68
##                                                        avg       = 130,800.46
##                                                        max       = 217,791.22
## Model F test:       Equal FMI                  F(   2,206760.5)  =       1.24
## Within VCE type:          OIM                  Prob > F          =     0.2891
##
## ------------------------------------------------------------------------------
##            e | Coefficient  Std. err.      t    P>|t|     [95% conf. interval]
## -------------+----------------------------------------------------------------
##         hist |
##     lobular  |    .107236   .1638183     0.65   0.513    -.2138461    .4283182
##       other  |   .1973595   .1287688     1.53   0.125    -.0550241     .449743
##              |
##        _cons |  -5.539115   .0631472   -87.72   0.000    -5.662883   -5.415346
##        ln(t) |          1  (exposure)
## ------------------------------------------------------------------------------
##
## . lincom _cons, eform
##
##  ( 1)  [e]_cons = 0
##
## ------------------------------------------------------------------------------
##            e |     exp(b)   Std. err.      z    P>|z|     [95% conf. interval]
## -------------+----------------------------------------------------------------
##          (1) |     .00393   .0002482   -87.72   0.000     .0034725    .0044478
## ------------------------------------------------------------------------------
##
## . lincom _cons + 2.hist, eform
##
##  ( 1)  [e]2.hist + [e]_cons = 0
##
```

```
## ---------------------------------------------------------------------------------
##            e |     exp(b)   Std. err.      z    P>|z|     [95% conf. interval]
## -------------+-------------------------------------------------------------------
##          (1) |   .0043749   .0006614   -35.93   0.000      .003253    .0058837
## ---------------------------------------------------------------------------------
##
## . lincom _cons + 3.hist, eform
##
##  ( 1)  [e]3.hist + [e]_cons = 0
##
## ---------------------------------------------------------------------------------
##            e |     exp(b)   Std. err.      z    P>|z|     [95% conf. interval]
## -------------+-------------------------------------------------------------------
##          (1) |   .0047875   .0005381   -47.53   0.000      .003841    .0059672
## ---------------------------------------------------------------------------------
##
## .
```

### Computational Details

- R version: R version 4.5.1 (2025-06-13)
- Base packages: stats, graphics, grDevices, utils, datasets, methods, base
- Other packages: multcomp 1.4.28, TH.data 1.1.3, MASS 7.3.65, mvtnorm 1.3.3, mice 3.17.0, kableExtra 1.4.0, broom 1.0.8, patchwork 1.3.0, survival 3.8.3, gtsummary 2.2.0, readxl 1.4.5, lubridate 1.9.4, forcats 1.0.0, stringr 1.5.1, dplyr 1.1.4, purrr 1.0.4, readr 2.1.5, tidyr 1.3.1, tibble 3.3.0, ggplot2 3.5.2, tidyverse 2.0.0, knitr 1.50

This document was generated on 2025-09-22 at 17:29.
